# Bayesian Adaptive Clinical Trials for Anti-Infective Therapeutics during Epidemic Outbreaks

**DOI:** 10.1101/2020.04.09.20059634

**Authors:** Qingyang Xu, Shomesh Chaudhuri, Danying Xiao, Andrew W. Lo

## Abstract

In the midst of epidemics such as COVID-19, therapeutic candidates are unlikely to be able to complete the usual multi-year clinical trial and regulatory approval process within the course of an outbreak. We apply a Bayesian adaptive patient-centered model—which minimizes the expected harm of false positives and false negatives—to optimize the clinical trial development path during such outbreaks. When the epidemic is more infectious and fatal, the Bayesian-optimal sample size in the clinical trial is lower and the optimal statistical significance level is higher. For COVID-19 (assuming a static *R*_0_ = 2 and initial infection percentage of 0.1%), the optimal significance level is 7.1% for a clinical trial of a non-vaccine anti-infective therapeutic clinical trial and 13.6% for that of a vaccine. For a dynamic *R*_0_ ranging from 2 to 4, the corresponding values are 14.4% and 26.4%, respectively. Our results illustrate the importance of adapting the clinical trial design and the regulatory approval process to the specific parameters and stage of the epidemic.

## 1 Introduction

With growing public concern over the outbreak of Coronavirus Disease 2019 (COVID-19), significant efforts have been undertaken by global biomedical stakeholders to develop effective diagnostics, vaccines, anti-viral drugs, medical devices, and other therapeutics against this highly infectious and deadly pandemic. While in the past, the traditional randomized clinical trial (RCT) and regulatory approval process often took several years (U.S. Food & Drug Administration, 2018)—longer than the typical duration of an epidemic outbreak (Pronker *et al*. 2013)—recently the FDA has responded with actions such as the Breakthrough Devices Program, Emergency Use Authorization (EUA) authority, and Immediately in Effect guidance documents to prevent novel diagnostics and therapeutics from lagging behind the urgent needs of the population. In this paper, we propose adapting yet another tool that the FDA has already been exploring for medical devices (Chaudhuri et al. 2018, 2019) to therapeutics for treating COVID-19 that are currently under development.

In recent years, Bayesian adaptive RCT protocols have been increasingly used to expedite the clinical trial process of potentially transformative therapies for diseases with high mortality rates (Berry, 2015). Currently, these protocols have mainly been applied within the oncology domain, such as I-SPY for breast cancer (Barker et al., 2009) and GBM AGILE for glioblastoma (Alexander *et al*. 2018). These studies use Bayesian inference algorithms to greatly reduce the number of patients needed to assess the therapeutic effects of a drug candidate, without lowering the statistical power of the final approval decision, as measured by Type I and II error rates. As a result, therapeutic candidates can progress more quickly through the regulatory process and reach patients faster and at lower cost.

For severe diseases with no curative treatments, such as pancreatic cancer, patients tend to tolerate a higher Type I error of accepting an ineffective therapy in exchange for a lower Type II error of rejecting an effective therapy as well as expedited approvals of potentially effective treatments. Based on this observation, a patient-centered Bayesian protocol was proposed (Isakov *et al*., 2018; Montazerhodjat *et al*., 2017) that incorporates patient values into clinical trial design and identify the optimal balance between the possibilities of false positives (Type I error) and false negatives (Type II error). For more severe diseases, this protocol sets a tolerated Type I error rate much larger than the traditional 5% threshold, which leads to higher rates of approvals and expedited approval decisions.

However, the original Bayesian adaptive RCT framework does not take into account patient risk preferences. To address this gap, Chaudhuri and Lo (2018) developed an adaptive version of the Bayesian patient-centered model that achieves an optimal balance between Type I and Type II error rates, significantly reducing the number of subjects needed in trials to achieve a statistically significant conclusion. A key feature of this model is the time evolution of the loss function of the Bayesian decision algorithm. This mechanism favors the expedited approval of diagnostic or therapeutic candidates that show early positive effects, since patients place a lower value on delayed approval of an effective diagnostic or therapy.

There is a natural but subtle analog to this dilemma in the case of therapeutics for an infectious disease during the course of an epidemic outbreak. Approving an effective therapeutic early will prevent future infections and deaths, while approving it later will save fewer people from infection. On the other hand, approving an ineffective therapeutic early will not prevent any future casualties. Worse still, it may prevent people from taking adequate precautions against infection, since they will falsely believe that they are safe from the disease after the advent of the ineffective therapy.

Moreover, the cost of Type I versus Type II error can differ from therapy to therapy. A novel vaccine that could trigger a significant immune response such as a cytokine storm has a much higher cost of a Type I error than a medical device such as an air filtration system designed to destroy virions through intense ultraviolet light. Therefore, the appropriate statistical threshold for approval should depend on the specific therapy, as well as the circumstances of the current burden of disease.

In this article, we apply the Bayesian adaptive protocol to anti-infective therapeutic development using a loss function that evolves over the course of an epidemic outbreak. We achieve an optimal balance between Type I and Type II errors for therapeutics that treat infectious diseases and identify the optimal time to reach the approval decision based on the accumulation of clinical evidence. Our results show that when the epidemic is more infectious, the necessary sample size of the RCT decreases, while the tolerable Type I error increases. This confirms our earlier intuition that potentially effective therapies that are known to be safe should receive expedited approval when an epidemic is spreading rapidly.

## 2 Multi-Group SEIR Epidemic Model

The starting point for our analysis is the Susceptible-Exposure-Infective-Removed (SEIR) epidemic model, which has been applied to model the outbreak of COVID-19 in China in a number of recent studies (Yang et al. 2020; Wu *et al*. 2020). The population of N subjects is partitioned into four distinct groups: susceptible (S), exposed (E), infectious (I), and removed (R). The time evolution of the epidemic is specified by the following group of ordinary differential equations:

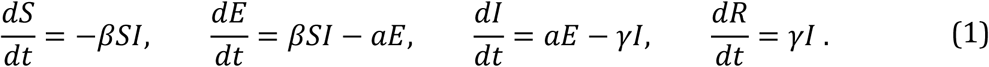

Here we use the convention that *S*(*t*), *E*(*t*), *I*(*t*), and *R*(*t*) are the proportions of the susceptible, exposed, infectious and removed populations, respectively, satisfying the conservation constraint for all *t*:

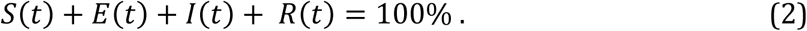

The parameters *β, a*, and *γ* denote the average rates of infection, incubation and recovery, respectively, and *μ* ∈ (0%, 100%) denotes the mortality rate of the epidemic. For example, if *μ* = 5%, we expect 5% of infected subjects will die from the disease. At time *t, μR*(*t*)*N* subjects will have died, and (1 - *μ*)*R*(*t*)*N* will have recovered.

A critical measure of the infectivity of an epidemic is its basic reproduction number, defined as *R*_0_ = *β*/*γ* in the SEIR model. This is the expected number of secondary infections caused by each infected subject in a population with no public health measures (such as quarantine, social-distancing, or vaccination).

A number of studies have used different statistical schemes to estimate *R*_0_ for COVID-19 during its initial outbreak period in central China in January 2020. These estimated values of *R*_0_ range from 2.2 (95% CI, 1.4 to 3.9) (Li *et al*., 2020) to 3.58 (95% CI, 2.89 to 4.39) (Zhao *et al*., 2020). Given the large uncertainty in the value of *R*_0_, we simulate therapeutic development under scenarios with constant *R*_0_ values of 2 and 4.

In addition, to model the impact of governmental nonpharmaceutical interventions (NPIs) on containing the spread of the epidemic, we consider a dynamic transmission SEIR model where the infection rate *β*(*t*) monotonically decreases in time as a result of the NPI. Specifically, we assume that *β*(*t*) takes the sigmoid functional form:

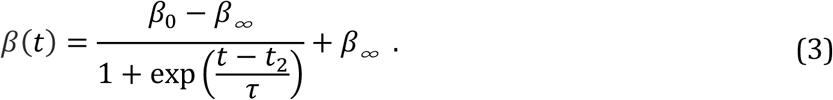

Here *β*_0_ and *β*_∞_ denote the infection rates in the initial and final stages of the epidemic (with *β*_0_ > *β*_∞_), respectively, *t*_2_ denotes the half-life of the decay in infection rate, and *τ* the length of the time window when this decay occurred. A larger difference *β*_0_ - *β*_∞_ corresponds to more significant reduction of epidemic transmission, a smaller value of *t*_2_ corresponds to a speedier decision to enforce the NPI, and a smaller value of *τ* corresponds to more strict enforcement of the NPI since *β*(*t*) decays more rapidly. We calibrate the values *β*_0_ = 3 and *β*_∞_ = 1.5 based on the estimates of the dynamic transmission rate of COVID-19 in Wuhan, China from December 2019 to February 2020 (Kucharski *et al*., 2020). We consider different values of *t*_2_ and *τ* to reflect the variability in timing and stringency of NPIs enforced by governments around the globe.

To model the significant variability in mortality rates of COVID-19 for patients in different age groups, we extend this basic model to a multi-group SEIR model, where the population is partitioned into five age groups, (1) below 49, (2) 50 to 59, (3) 60 to 69, (4) 70 to 79, and (5) above 80. We use *S*_*i*_, *E*_*i*_, *I*_*i*_, and *R*_*i*_ to denote the corresponding type in each group (and continue to use *S, E, I*, and *R* for the total proportion of each type in all groups). The dynamics of the epidemic are specified by the modified ordinary differential equations:

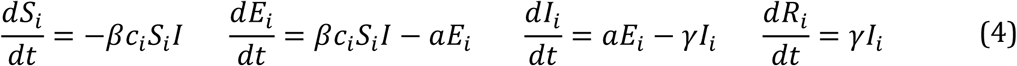

Here *c*_*i*_ denotes the contact rate of the susceptible subjects in the *i*^th^ age group with the total infected population *I* of all groups. This contact rate is measured relative to group 1, which we normalize to *c*_l_ = 1. In the case of COVID-19, although the mortality rate is much higher for senior populations (Onder *et al*., 2020), the elderly also tends to have less frequent contact with the infected population outside the household (Walker *et al*., 2020).

We solve the differential equations in the multi-group SEIR model using the ODE45 solver in MATLAB 2019a with initial conditions for each age group:

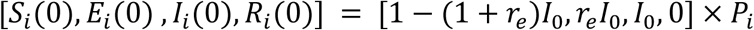

The parameter *I*_0_ denotes the proportion of the initially infected population, *r*_e_ is the ratio of initially exposed and infected subjects, and *P*_*i*_ is the percentage of the *i*^th^ age group in the population. The assumed demographic, contact rate, and mortality rate values are summarized in Table 2.

## 3 A Bayesian Patient-Centered Approval Process

Similar to Chaudhuri and Lo (2018), we develop a Bayesian patient-centered decision model for RCT approval which minimizes the expected loss (or harm) incurred on the patients by optimally balancing the losses of Type I and Type II errors. Here the loss does not refer to financial costs afforded by the patients, but rather the loss in patient value (i.e. how much patients weigh the relative harms of infection and death). We assign the losses per patient of being susceptible, infected, and deceased. Since Bayesian decision thresholds are invariant under the rescaling of the losses, we normalize by setting the loss per patient infection *L*_*i*_ = 1. We then assign the loss per patient death relative to *L*_*i*_ as *L*_D_, and the loss due to susceptibility to the disease as *L*_S_. The parameter values we assume, summarized in Table 1, are meant to represent one reasonable valuation of the relative losses. However, in practice patient value will differ from one patient group another, especially given the large variability of mortality rate of COVID-19 in different age groups (Onder *et al*., 2020). Here we report the main results of optimal sample size and statistical significance (Table 3 and 4) assuming *L*_D_ = 100. The results for *L*_D_ = 10 are provided in Supplementary Materials.

**Table 1.** Simulation parameters and values.

| Parameter | Description (Source) | Value(s) |
| --- | --- | --- |
| $R_0$ | Basic reproduction number<br>(Li *et al.* 2020; Zhao *et al.*, 2020) | 2, 4 |
| $a$ | Incubation rate (per week)<br>(Yang *et al.* 2020) | 1 |
| $\gamma$ | Recovery rate (per week)<br>(Yang *et al.* 2020) | 1 |
| $I_0$ | Initial proportion of infected population | 0.1%, 0.01% |
| $r_e$ | Ratio of initially exposed and infected populations | 10 |
| $[\beta_0, \beta_\infty]$ | Initial and final infection rate in the dynamic transmission model (Kucharski *et al.* 2020) | [3, 1.5] |
| $t_2$ | Half-life of decay in the dynamic model (week) | 3, 6 |
| $\tau$ | Window length of decay in the dynamic model (week) | 0.5, 1 |
| $N$ | Population size (million) | 300 |
| $\kappa$ | Weekly subject enrollment in each arm of RCT | 100 |
| $p_0^{th}$ | Prior probability of having an ineffective anti-infective therapeutic (Wong *et al.*, 2019) | 77% |
| $p_0^{vac}$ | Prior probability of having an ineffective vaccine (Wong *et al.*, 2019) | 60% |
| $\Delta t$ | Time needed to assess the efficacy of the treatment (week) | 1 |
| $\rho$ | Signal to noise ratio ($\delta/\sigma$) of treatment effect (Chaudhuri and Lo, 2018) | 0.25 |
| $\text{Power}_{\max}$ | Maximum power of Bayesian decision model (Isakov *et al.*, 2019) | 0.9 |
| $L_D$ | Loss per capita from death by infection | 10, 100 |
| $L_I$ | Loss per capita from being infected | 1 |
| $L_S$ | Loss per capita from being susceptible without precaution | 0.2 |

**Table 2.**
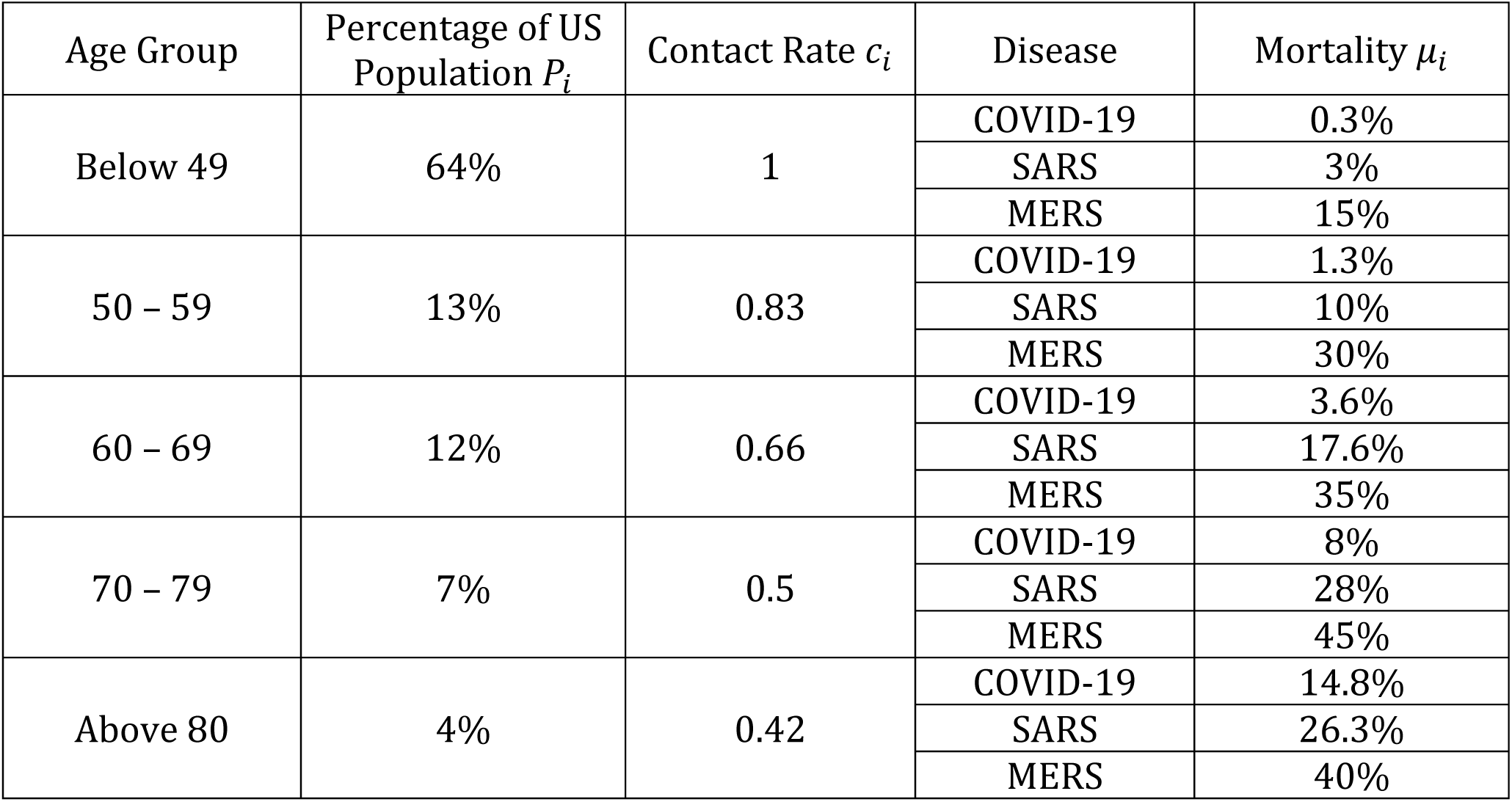
Demographic (US Census Bureau, 2018), relative contact rate (Walker *et al*. 2020) and mortality (Onder *et al*. 2020; World Health Organization 2003; 2019; 2020) profile of various age groups for COVID-19, SARS and MERS.

| Age Group | Percentage of US Population $P_i$ | Contact Rate $c_i$ | Disease | Mortality $\mu_i$ |
| --- | --- | --- | --- | --- |
| Below 49 | 64% | 1 | COVID-19 | 0.3% |
|  |  |  | SARS | 3% |
|  |  |  | MERS | 15% |
| 50 – 59 | 13% | 0.83 | COVID-19 | 1.3% |
|  |  |  | SARS | 10% |
|  |  |  | MERS | 30% |
| 60 – 69 | 12% | 0.66 | COVID-19 | 3.6% |
|  |  |  | SARS | 17.6% |
|  |  |  | MERS | 35% |
| 70 – 79 | 7% | 0.5 | COVID-19 | 8% |
|  |  |  | SARS | 28% |
|  |  |  | MERS | 45% |
| Above 80 | 4% | 0.42 | COVID-19 | 14.8% |
|  |  |  | SARS | 26.3% |
|  |  |  | MERS | 40% |

**Table 3.** Simulation results of a Bayesian adaptive RCT on non-vaccine anti-infective therapeutics obtained from 10,000 Monte Carlo runs and assuming *L*_-*D*_ = 100. *R*_0_ denotes the basic reproduction number, *µ* the disease morality, and *I*_0_ the proportion of initial infected subjects. Sample size refers to the number of subjects enrolled in each arm of the RCT. SD denotes standard deviation, and IQR the interquartile range about the median. *R*_0_(t) denotes the use of a dynamic transmission model with half-life *t*_2_ = 3 weeks and window length τ = 1 week.

| Epidemic Parameters |  |  | Non-Adaptive |  | Power % | Adaptive (100,000 Monte Carlo Runs) |  |  |  |  |  |
| --- | --- | --- | --- | --- | --- | --- | --- | --- | --- | --- | --- |
| | | | | | | Sample Size (H=0) | | Sample Size (H=1) | | $\alpha$ % | Power % |
| $R_0$ | $\mu$ | $I_0$ | Sample Size | $\alpha$ % | | Mean (SD) | Median (IQR) | Mean (SD) | Median (IQR) | | |
| 2 | COVID-19 | 0.1% | 242 | 7.1 | 90 | 135 (103) | 105 (63, 176) | 148 (107) | 119 (73, 192) | 5.8 | 91.5 |
| 4 | COVID-19 | 0.1% | 158 | 17.3 | 90 | 115 (83) | 91 (56, 149) | 98 (80) | 74 (42, 128) | 14.4 | 92.1 |
| $R_0(t)$ | COVID-19 | 0.1% | 176 | 14.4 | 90 | 118 (85) | 95 (57, 153) | 108 (85) | 84 (49, 140) | 11.7 | 92.2 |
| 2 | COVID-19 | 0.01% | 399 | 1.2 | 90 | 150 (128) | 110 (64, 191) | 248 (154) | 211 (139, 317) | 1.0 | 91.0 |
| 4 | COVID-19 | 0.01% | 274 | 5.0 | 90 | 140 (110) | 106 (64, 180) | 168 (116) | 136 (86, 216) | 4.1 | 91.4 |
| $R_0(t)$ | COVID-19 | 0.01% | 304 | 3.6 | 90 | 145 (119) | 108 (64, 187) | 184 (120) | 153 (97, 239) | 3.0 | 91.2 |
| 2 | SARS | 0.1% | 164 | 16.3 | 90 | 117 (85) | 94 (57, 150) | 101 (81) | 77 (45, 132) | 13.9 | 92.3 |
| 4 | SARS | 0.1% | 112 | 27.8 | 90 | 98 (72) | 79 (47, 128) | 72 (64) | 51 (27, 95) | 23.3 | 93.2 |
| $R_0(t)$ | SARS | 0.1% | 107 | 29.2 | 90 | 96 (71) | 78 (45, 126) | 70 (64) | 50 (26, 92) | 25.1 | 93.4 |
| 2 | MERS | 0.1% | 88 | 35.3 | 90 | 87 (66) | 70 (40, 115) | 59 (57) | 39 (20, 77) | 29.8 | 93.7 |
| 4 | MERS | 0.1% | 63 | 45.2 | 90 | 73 (59) | 59 (30, 100) | 46 (49) | 28 (14, 60) | 38.8 | 94.5 |
| $R_0(t)$ | MERS | 0.1% | 44 | 54.3 | 90 | 61 (54) | 48 (20, 86) | 36 (43) | 19 (9, 46) | 47.0 | 94.8 |

**Table 4.** Simulation results of Bayesian adaptive RCT for vaccines obtained from 10,000 Monte Carlo runs and assuming *L*_*D*_ = 100. *R*_0_ denotes the basic reproduction number, *µ* the disease morality, and *I*_0_ the proportion of initial infected subjects. Sample size refers to the number of subjects enrolled in each arm of the RCT. SD denotes standard deviation, and IQR the interquartile range about the median. *R*_0_(t) denotes the use of a dynamic transmission model with half-life *t*_2_ = 3 weeks and window length τ = 1 week.

| Epidemic Parameters |  |  | Non-Adaptive |  | Power % | Adaptive (100,000 Monte Carlo Runs) |  |  |  |  |  |
| --- | --- | --- | --- | --- | --- | --- | --- | --- | --- | --- | --- |
| | | | | | | Sample Size (H=0) | | Sample Size (H=1) | | $\alpha$ % | Power % |
| $R_0$ | $\mu$ | $I_0$ | Sample Size | $\alpha$ % | | Mean (SD) | Median (IQR) | Mean (SD) | Median (IQR) | | |
| 2 | COVID-19 | 0.1% | 181 | 13.6 | 90 | 122 (91) | 95 (58, 158) | 112 (87) | 86 (51, 145) | 11.3 | 92.4 |
| 4 | COVID-19 | 0.1% | 111 | 28.1 | 90 | 97 (71) | 78 (47, 127) | 72 (64) | 52 (27, 95) | 23.4 | 92.8 |
| $R_0(t)$ | COVID-19 | 0.1% | 117 | 26.4 | 90 | 71 (49) | 80 (49, 132) | 74 (67) | 53 (29, 98) | 21.8 | 93.1 |
| 2 | COVID-19 | 0.01% | 342 | 2.3 | 90 | 148 (124) | 110 (64, 191) | 212 (137) | 177 (115, 275) | 2.1 | 90.9 |
| 4 | COVID-19 | 0.01% | 232 | 7.9 | 90 | 132 (100) | 104 (61, 171) | 142 (103) | 113 (69, 184) | 6.6 | 91.4 |
| $R_0(t)$ | COVID-19 | 0.01% | 244 | 6.9 | 90 | 132 (101) | 102 (63, 171) | 148 (106) | 118 (73, 191) | 5.4 | 91.7 |
| 2 | SARS | 0.1% | 99 | 31.7 | 90 | 91 (67) | 74 (44, 119) | 65 (62) | 44 (24, 86) | 26.6 | 93.5 |
| 4 | SARS | 0.1% | 65 | 44.3 | 90 | 74 (59) | 60 (32, 99) | 47 (50) | 29 (14, 61) | 37.6 | 94.5 |
| $R_0(t)$ | SARS | 0.1% | 50 | 51.3 | 90 | 65 (55) | 52 (23, 91) | 40 (45) | 23 (11, 51) | 44.2 | 94.8 |
| 2 | MERS | 0.1% | 27 | 64.2 | 90 | 49 (49) | 36 (11, 71) | 28 (37) | 13 (6, 34) | 55.4 | 95.9 |
| 4 | MERS | 0.1% | 21 | 68.1 | 90 | 45 (48) | 31 (8, 66) | 25 (35) | 11 (5, 29) | 58.9 | 96.3 |
| $R_0(t)$ | MERS | 0.1% | 7 | 79.2 | 90 | 33 (41) | 14 (4, 50) | 17 (27) | 6 (3, 18) | 69.1 | 97.2 |

We simulate the multi-group SEIR model over a time period of *T* weeks, where *T* is the duration of the epidemic outbreak. Let *κ* denote the weekly subject enrollment rate in each arm of the clinical trial. We assume that the value of *R*_0_ is known (or well-estimated) at initial time *t* = 0 and stays constant during the course of the outbreak. At time *t* ∈ [0, *T*], the Bayesian loss C_ij_(t) of choosing the action *Ĥ* = *i* under *H* = *j* is defined as:

| | $\hat{H} = 0$ (do not approve) | $\hat{H} = 1$ (approve) |
| --- | --- | --- |
| $H = 0$<br>(no effect) | 0 | $(S(t) - S(T))NL_S$ |
| $H = 1$<br>(effective) | $R(T)N(L_I + \mu L_D)$ | $CI(t)NL_I + \mu R(t)NL_D$ |

where we define the cumulative number of infected patients *CI*(*t*) until time t:

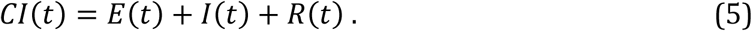

By design, this loss function penalizes Type I errors early in the epidemic by the susceptible term, (*S*(*t*) - *S*(*T*))*NL*_S_. We subtract the base level *S*(*T*) from *S*(*t*) since the multi-group SEIR model predicts that *S*(*T*)*N* subjects will not be infected by the epidemic. A Type I error at an earlier time will expose more currently susceptible population to the epidemic, since they will falsely believe that they are safe from the disease after the advent of the ineffective therapeutic. On the other hand, the loss function also penalizes *correct* approval decisions made at later stages of an epidemic via to the cumulative infected and death terms, *CI*(*t*) and *μR*(*t*). A correct but delayed approval decision for the therapeutic is less valuable since it will save fewer susceptible people from infection and death.

The Bayesian decision model considers the null hypothesis *H* = 0 that the anti-infective therapeutic (or vaccine) has no clinical effect, against the alternative hypothesis that it has positive clinical effect with signal-to-noise ratio *ρ* (Chaudhuri and Lo, 2018). We use *p*_0_and *p*_l_ to denote the Bayesian prior probabilities of *H* = 0 and *H* = 1, respectively.

This patient-value model imposes higher losses for incorrect approvals at earlier stages and correct approvals at later stages of an epidemic. Under these constraints, the Bayesian decision algorithm yields the sample size and statistical significance threshold of the RCT that optimally balances Type I and Type II error.

## 4 Results

We simulate an epidemic outbreak over a time period of *T* weeks, where *T* is the duration of the outbreak. For an epidemic with higher infectivity, its duration is shorter, which puts more pressure to reach a timely approval decision. To avoid numerical instability, we formally define *T* as the first time when the number of cumulative infected patients reaches 99.9% of total infections predicted by the SEIR model. We assume an age-specific mortality rate *μ* at the level of COVID-19 (Onder *et al*. 2020; World Health Organization, 2020), and incubation and recovery periods of 7 days each (Yang *et al*. 2020). These estimated parameters can all be challenged to varying degrees, depending on the specific drug-indication pair under consideration and the particular circumstances of the epidemic, but they are meant to be representative for a typical anti-infective therapeutic during the midst of a growing epidemic.

We also assume that it takes 7 days after injection to assess the efficacy of the therapeutic on each subject. We adopt the optimization scheme of Montazerhodjat *et al*. (2017) to find the optimal Type I and Type II error rates of the non-adaptive Bayesian RCT. To represent typical practice of the pharmaceutical industry, we optimize under the upper bound on the model’s power Power_max_ = 90% (Isakov *et al*. 2019). We then use these optimal error rates as our stopping criteria to simulate the sequential decision process of a Bayesian adaptive RCT via Monte Carlo simulation (Chaudhuri and Lo, 2018). The simulation results are summarized in Table 3.

We separate the results into two distinct types of therapeutics—non-vaccine anti-infectives and vaccines—because of the differences in their historical probabilities of success. Vaccine development programs have an estimated PoS of 40% as of 2019Q4 (https://projectalpha.mit.edu) whereas the corresponding figure for non-vaccine anti-infectives is 23% (Wong et al., 2020).

## 4.1 Non-Vaccine Anti-Infective Therapeutics

### Static Transmission Rate

We first analyze the case when the infectivity *R*_0_ remains constant in time (e.g. in the absence of effective NPIs). For the fixed-sample Bayesian RCT on a non-vaccine anti-infective therapeutic, as *R*_0_ increases from 2 to 4 (Rows 1 to 2 of Table 3), the optimal sample size of each experimental arm decreases from 242 to 152 and the optimal Type I error rate drastically increases from 7.1% to 17.3% (Figure 1), much higher than the traditional 5% threshold. As the epidemic spreads across the population more rapidly, the Bayesian RCT model has greater pressure to expedite the approval process and a much higher tolerance of false positive outcomes.

**Figure 1.**
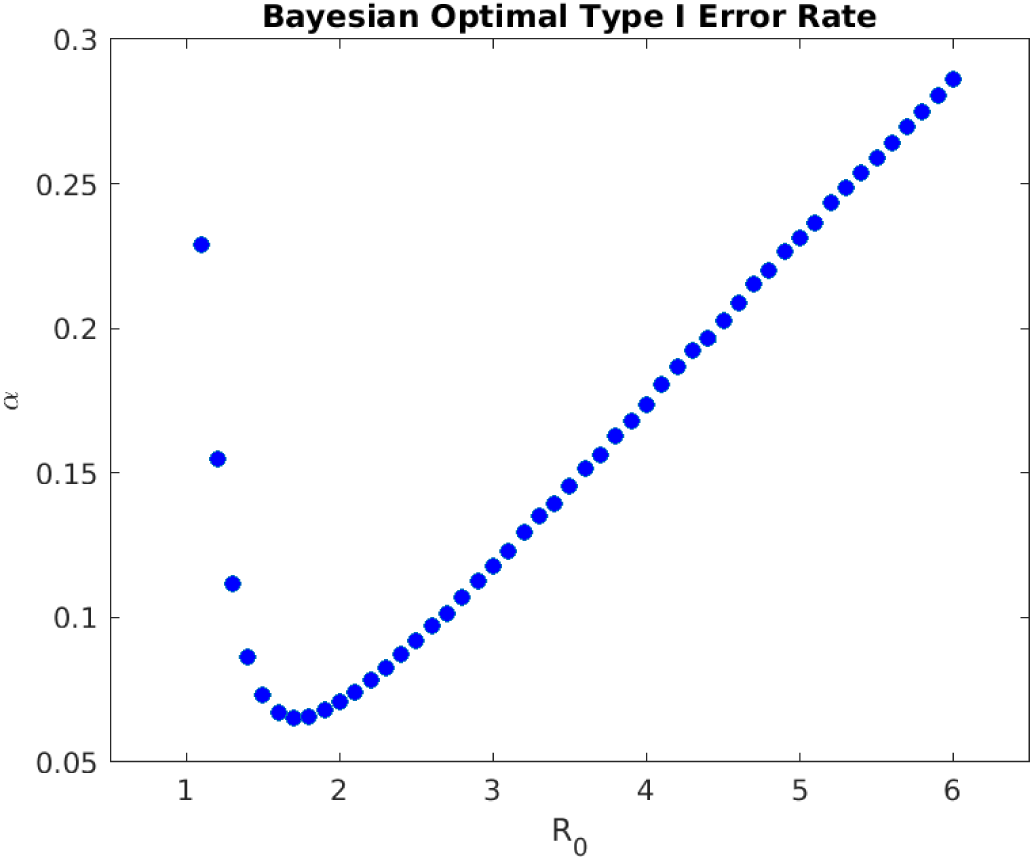
Optimal Type I error rate *α* of a non-adaptive Bayesian RCT vs. basic reproduction number *R*_0_ (assuming *I*_0_ = 0. 1%, *L*_*D*_ = 100, disease mortality of COVID- 19, and constant *R*_0_). The Bayesian decision model yields a higher *α* for epidemics with high and low infectivity.

For the Bayesian adaptive RCT, when the therapeutic is ineffective (*H* = 0), the average sample size required to reject the therapeutic is much smaller than that of the non-adaptive version (Columns 7 and 8 of Table 3). Also, the required sample size decreases with the infectivity *R*_0_ in both mean and quartiles, yet always achieves Type I error rate *α* below that of the non-adaptive version (Column 11). The adaptive Bayesian decision model is able to reject an ineffective therapeutic with a relatively small sample size and a bounded false-positive rate.

On the other hand, when the therapeutic is effective (*H* = 1), as *R*_0_ increases from 2 to 4, the average sample size required by the Bayesian adaptive RCT decreases from 148 to 78 (Columns 9 and 10 of Table 3). The Bayesian adaptive model places more weight on approving an effective therapeutic earlier to prevent future infections when the epidemic is more infectious. Despite the smaller sample size, the model still retains an empirical power above 91.0% for all values of *R*_0_ (Column 12). The Bayesian adaptive model simultaneously expedites the approval of an effective therapeutic and retains a bounded false-negative rate. The results are illustrated in Figure 2.

**Figure 2.**
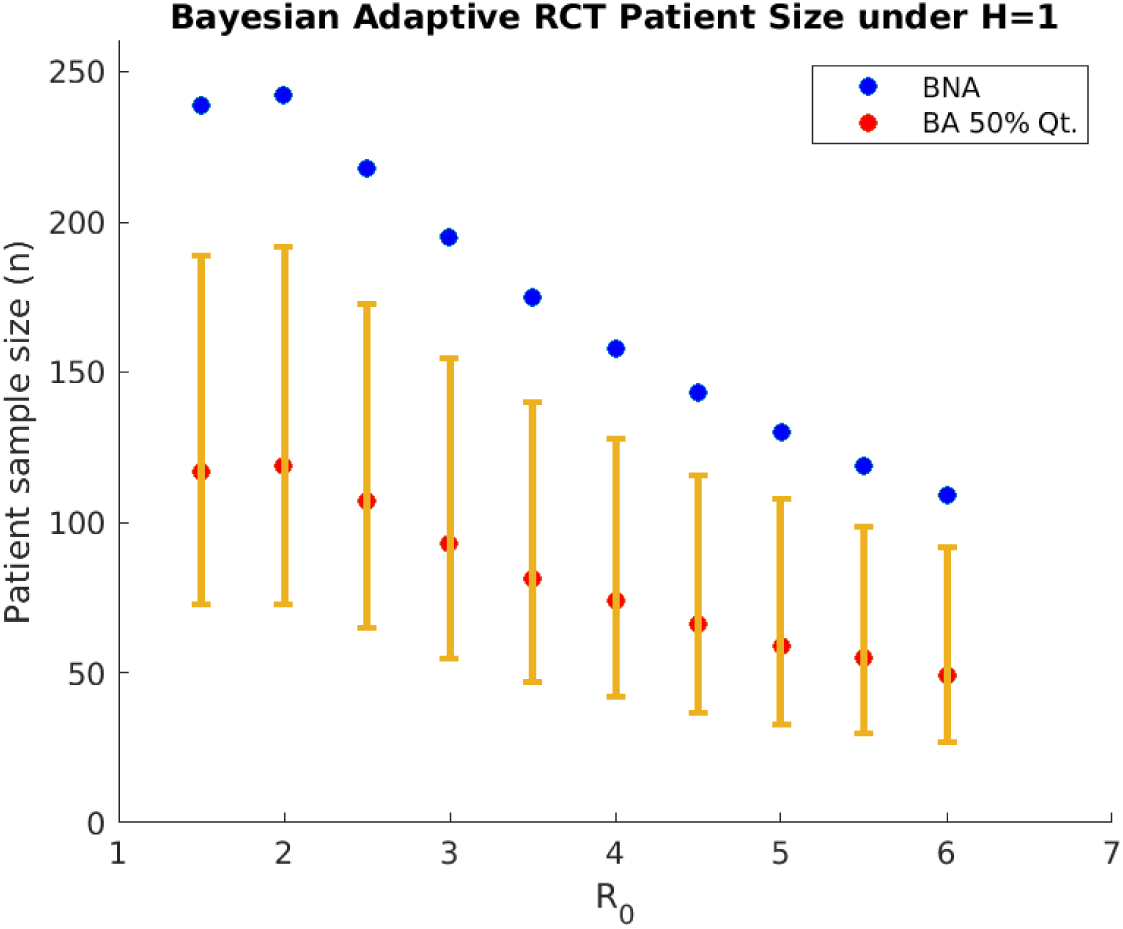
Subject sample size in each arm of a Bayesian adaptive RCT under H = 1 decreases with the basic reproduction number *R*_0_ (assuming *I*_0_ = 0. 1%, *L*_*D*_ = 100 and disease mortality of COVID-19). BNA denotes Bayesian non-adaptive optimal; BA 50% denotes median patient size of Bayesian adaptive. The 25% and 75% quantiles of Bayesian adaptive patient size are shown as lower and upper ends of the error bar.

Furthermore, as the proportion of the initially infected population *I*_0_ decreases from 0.1% to 0.01% (Rows 4 to 6 of Table 3), the optimal sample sizes for non-adaptive and adaptive RCTs both increase, while the optimal Type I error rates decrease. Beginning the clinical trials for a therapeutic during the earlier stages of an epidemic outbreak reduces the need to expedite the approval process in order to contain its future spread. Clinicians and researchers have more time to evaluate the efficacy of a therapeutic and record adverse effects by testing it on a larger number of subjects, which leads to a lower Type I error rate.

Finally, when the mortality rate *μ* increases from the level of COVID-19 (Onder *et al*. 2020; World Health Organization, 2020), to the level of SARS (World Health Organization, 2003), and further to the level MERS (World Health Organization, 2019), the optimal sample sizes for both non-adaptive and adaptive Bayesian models decrease and the optimal Type I error rates increase (Rows 7 to 12 of Table 3). When the epidemic is more lethal, the Bayesian adaptive model requires fewer subjects in the RCT, since both Type I and Type II errors will lead to greater losses due to death by infection. The higher death tolls provides significantly more incentive in the Bayesian adaptive framework to approve the therapeutic in the hopes of saving more people from future infection and death.

One interesting feature of the Bayesian decision model is that the optimal Type I error rate is not a monotonic function *R*_0_, but rather has a global minimum of 8% at *R*_0_ = 1.7 for COVID-19, as shown in Figure 1. As *R*_0_ decreases below 1.7, the optimal Type I error rate increases. The intuition for this result that we define the loss of Type I error as the excess risk of being susceptible to infection (*S*(*t*) - *S*(*T*))*NL*_S_, where *S*(*T*) is the fraction of the population that remains uninfected throughout the epidemic outbreak. When *R*_0_ is small, *S*(*T*) is close to 100% and the excess risk (*S*(*t*) - *S*(*T*))*NL*_S_ is small compared to the benefit of preventing future deaths. Therefore, when the epidemic is not very infectious, the Bayesian decision model expedites the approval decision. This also confirms the intuition that smaller sample sizes are required in adaptive trials for diseases that affect a small fraction of the population. If we instead define the loss of Type I error as the absolute risk of being susceptible *S*(*t*)*NL*_S_, we find that the optimal Type I error indeed monotonically increases with *R*_0_, as shown in Figure 11 of Supplementary Materials.

### Dynamic Transmission Rate

The results for the dynamic transmission model with *β*_0_ = 3, *β*_∞_ = 1.5, *t*_2_ = 3 weeks and *τ* = 1 week are summarized in Table 3. For COVID-19 (rows 3 and 6), we find that the Bayesian optimal sample size and Type I error rate of the dynamic transmission model lie in between the scenarios *R*_0_ = 2 and *R*_0_ = 4. This suggests that timely and effective government interventions will protect more subjects from infection and allow more time for the RCT.

However, for the more fatal SARS and MERS (rows 9 and 12), the dynamic transmission model sets higher optimal Type I error *α* and smaller sample size than *R*_0_ = 4. This is due to the U-shaped curve of optimal *α* vs. *R*_0_, shown in Figure 1. When the NPI reduces *R*_0_(*t*) below a certain threshold, the optimal *α* starts to increase. For highly fatal epidemics, when the government adopts NPIs to protect most of the susceptible population from infection, the regulatory priority should be to expedite potentially effective treatments that can help current patients since the loss of Type I error is much lower than that of the Type II error.

In addition, we investigate the impact of the timing and stringency of NPIs enforced by the government with different values of *t*_2_ and *τ*. The results are summarized in Table 5. We find that the optimal Type I error is larger for *t*_2_ = 3 weeks than *t*_2_ = 6 weeks. Therefore, if the government adopts well-enforced NPIs early on (such as the lockdown of Wuhan, China) to protect the susceptible population, this will reduce the loss associated with Type I error, leading to expedited approvals of potentially effective therapeutics. Furthermore, the sooner an effective therapeutic is approved, the sooner will NPIs be lifted.

**Table 5.** Optimal sample size and Type I error *α* of Bayesian non-adaptive RCT for non-vaccine anti-infective therapeutics for dynamic transmission model. *R*_0_ denotes the basic reproduction number, *µ* the disease morality, and *I*_0_ the proportion of initial infected subjects. Sample size refers to the number of subjects enrolled in each arm of the RCT. *R*_0_(t) denotes the use of a dynamic transmission model with *β*_0_ = 3, *β*_∞_ = 1. 5.

| Disease | $I_0$ | $R_0$ | $t_2$(week) | $\tau$ (week) | Sample Size | $\alpha$ % | Power % |
| --- | --- | --- | --- | --- | --- | --- | --- |
| COVID-19 | 0.1% | 2 | NA | NA | 242 | 7.1 | 90 |
|  |  | 4 | NA | NA | 158 | 17.3 | 90 |
| | | $R_0(t)$ | 3 | 0.5 | 166 | 16.0 | 90 |
| | | $R_0(t)$ | 3 | 1 | 176 | 14.4 | 90 |
| | | $R_0(t)$ | 6 | 0.5 | 176 | 14.4 | 90 |
| | | $R_0(t)$ | 6 | 1 | 177 | 14.2 | 90 |
| SARS | 0.1% | 2 | NA | NA | 164 | 16.3 | 90 |
|  |  | 4 | NA | NA | 112 | 27.8 | 90 |
| | | $R_0(t)$ | 3 | 0.5 | 100 | 31.3 | 90 |
| | | $R_0(t)$ | 3 | 1 | 107 | 29.2 | 90 |
| | | $R_0(t)$ | 6 | 0.5 | 118 | 26.2 | 90 |
| | | $R_0(t)$ | 6 | 1 | 119 | 25.9 | 90 |
| MERS | 0.1% | 2 | NA | NA | 88 | 35.3 | 90 |
|  |  | 4 | NA | NA | 63 | 45.2 | 90 |
| | | $R_0(t)$ | 3 | 0.5 | 41 | 55.9 | 90 |
| | | $R_0(t)$ | 3 | 1 | 44 | 54.3 | 90 |
| | | $R_0(t)$ | 6 | 0.5 | 59 | 47.0 | 90 |
| | | $R_0(t)$ | 6 | 1 | 60 | 46.5 | 90 |

### 4.2 Vaccines

We repeat the above analysis for RCT of vaccines using a prior probability of having an effective vaccine 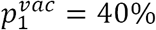 as reported at https://projectalpha.mit.edu for 2019Q4. The simulation results are summarized in Table 4. Overall, we observe the same pattern in the optimal sample size and Type I error rates on infectivity, mortality, and proportion of initial infections. However, since *p*_l_ is higher for vaccines, the Bayesian decision model requires fewer subjects on average in the RCT to ascertain the positive effects of the vaccine, compared to the case of anti-infective therapeutics in Table 3. We find that vaccines should receive even more expedited evaluation.

To assess the robustness of our model’s predictions against the assumed values of model parameters, we perform a sensitivity analysis on the parameter values. The results are summarized in Supplementary Materials.

## 5 Discussion

A natural consequence of using a patient-centered framework for determining the approval threshold is, of course, more false positives—and the potential for a greater number of patients with adverse side-effects—in cases where the burden of disease is high. These false positives can be addressed through more vigilant post-approval surveillance by regulatory agencies and greater requirements for drug and device companies to provide such patient-level data to the regulator following approval. Failure to provide such data or evidence of an ineffective therapy can be grounds for revoking the approval.

However, past experience shows that withdrawing an approved drug can be challenging and disruptive for several reasons (Onakpoya, Heneghan, and Aronson, 2016). Therefore, implementing the patient-centered approach may require creating a new category of temporary approvals for crisis situations involving urgent needs at national or international levels, similar to the FDA’s EUA program. Such a program might involve provisional approval of a candidate therapy consisting of a one- or two-year license—depending on the nature of the drug-indication pair—to market the therapy to a pre-specified patient population, no off-label use of the therapy, and regular monitoring and data reporting to the regulator by the manufacturer and/or patients’ physicians during the licensing period (Lo, 2017). At the end of this trial period, one of two outcomes would occur, depending on the accumulated data during this period: (a) the “urgent needs” license expires; or (b) the license converts to the traditional regulatory license. Of course, at any point during the trial period, the regulator can terminate the license if the data show that the therapeutic is ineffective and/or unsafe.

While such a process may impose greater burdens on patients, manufacturers, and regulators, it may still be worthwhile if it brings faster or greater relief to patients facing mortal illnesses and extreme suffering. In this respect, an urgent-needs program may be viewed as a middle ground between a standard clinical trial and an approval, similar in spirit to the adaptive designs of sophisticated clinical trials with master protocols such as I-SPY 2, LUNG-MAP, and GBM-AGILE, in which patient care and clinical investigations are simultaneously accomplished. Also, because the Centers for Medicare and Medicaid Services (CMS) has demonstrated a willingness to cover the cost of certain therapeutics for which evidence is still being generated (see, for example, CMS’s “coverage with evidence” programs listed at https://go.cms.gov/2v6ZxWm), additional economic incentives may be available to support such temporary licenses.

## 6 Conclusion

We apply the Bayesian adaptive patient-centered model of Chaudhuri and Lo (2018) to clinical trials for therapeutics that treat infectious diseases during an epidemic outbreak. Using a simple epidemiological model, we find that the optimal sample size in the clinical trial decreases with the infectivity of the epidemic, measured by the basic reproduction number *R*_0_. At the same time, the optimal Type I error rate increases with *R*_0_. Lower levels of initial infection increase the number of subjects required to verify the therapeutic efficacy of the therapeutic under investigation, while higher levels of mortality increase the optimal sample size. The results confirm our intuition that clinical trials should be expedited and a higher false positive rate should be tolerated when the epidemic spreads more rapidly through the population, has a higher mortality rate, and has already infected a sizable portion of the population at the beginning of the RCT.

To provide transparency for how a patient-centered approach differs from the traditional statistical framework in the anti-infectives context, we used a relatively simple mathematical model of epidemic disease dynamics to estimate the societal loss in an outbreak. More sophisticated epidemiological models can easily be incorporated into our framework at the cost of computational tractability and transparency.

One interesting trade-off to be explored is the difference between COVID-19 vaccine and an anti-viral treatment that can cure an infected patient. While prevention through vaccination is the ultimate solution, a successful treatment for the disease using repurposed drugs that have already been approved for other indications (and whose safety profile has already been established) may be even more valuable, especially if it can be deployed in the nearer term and reduce the growing fear and panic among the general population. In such cases, the approval threshold should clearly reflect these cost/benefit differences.

Of course, in practice regulators consider many factors beyond *p*-values in making its decisions. However, that process is opaque even to industry insiders, and the role of patient preferences is unclear. The proposed patient-centered approach provides a systematic, objective, adaptive, and repeatable framework for explicitly incorporating patient preferences and burden-of-disease data in the therapeutic approval process. This framework also fulfills two mandates for the FDA, one from the fifth authorization of the Prescription Drug User Fee Act (PDUFA) for an enhanced quantitative approach to the benefit-risk assessment of new drugs (U.S FDA, 2013), and the other from Section 3002 of the 21^st^ Century Cures Act of 2016 requiring the FDA to develop guidelines for patient-focused drug development, which includes collecting patient preference and experience data and explicitly incorporating this information in the drug approval process.

We hope this work will shed further insight into improving the current clinical trial process for infectious disease therapeutics and contribute to the timely development of effective treatments for COVID-19 patients in particular.

## Data Availability

All data used in this project are commercially available to other authors on the same terms.

https://informa.com

## Supplementary Materials

**Figure 3.**
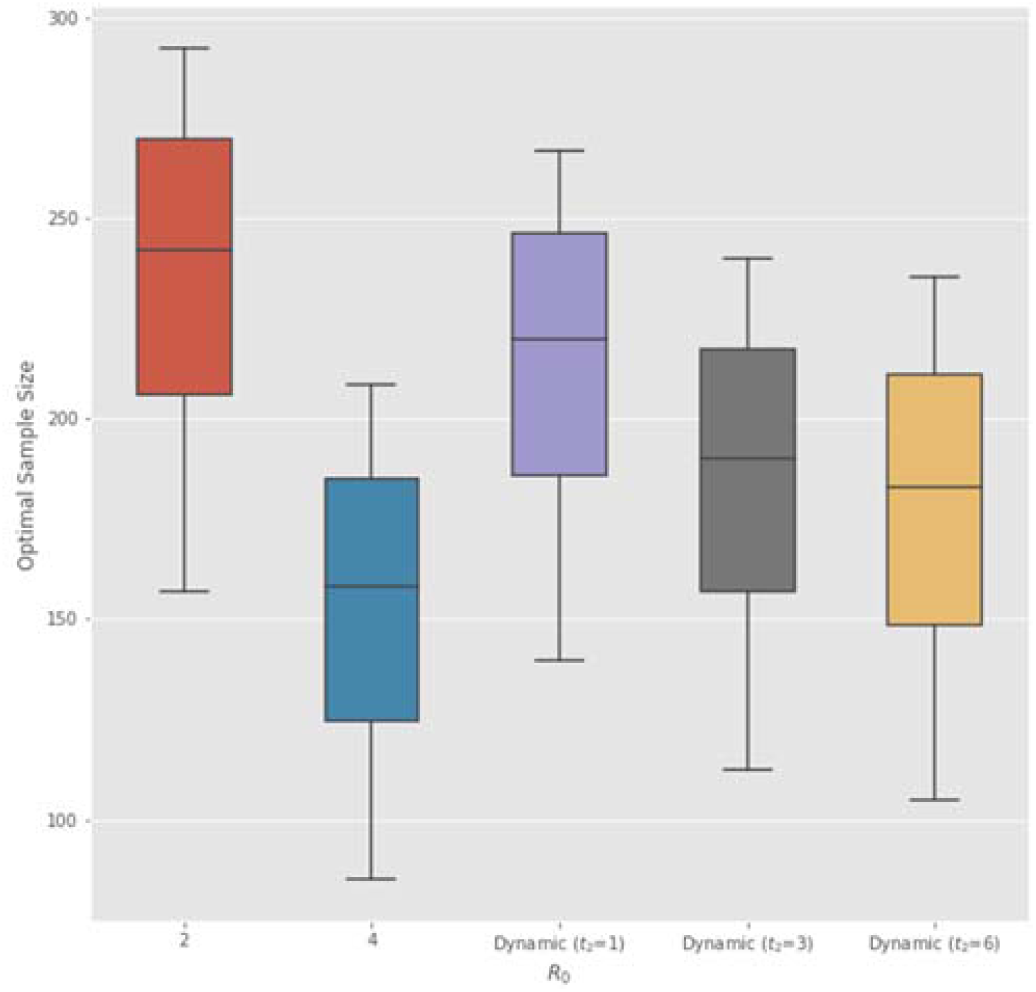
Sensitivity of optimal sample size (in each arm of non-adaptive Bayesian RCT) to the weekly subject enrollment rate *κ*. From the lower to upper end of each box plot, the five-parameter summary corresponds to [50%, 75%, 100%, 125%, 150%] of the baseline value *κ* = 100 used in our analysis (Table 1).

**Figure 4.**
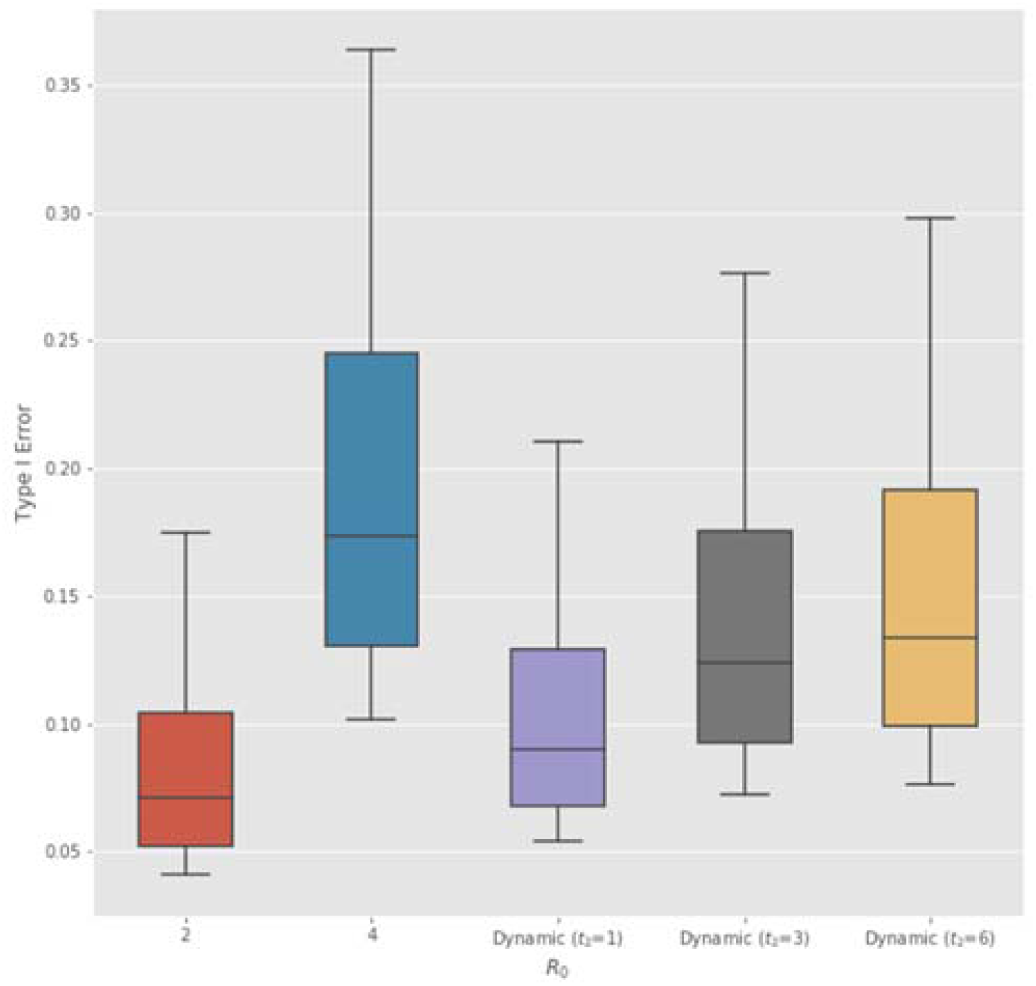
Sensitivity of optimal Type I error rate to the weekly subject enrollment rate *κ*. From the upper to lower end of each box plot, the five-parameter summary corresponds to [50%, 75%, 100%, 125%, 150%] of the baseline value *κ* = 100 used in our analysis (Table 1).

**Figure 5.**
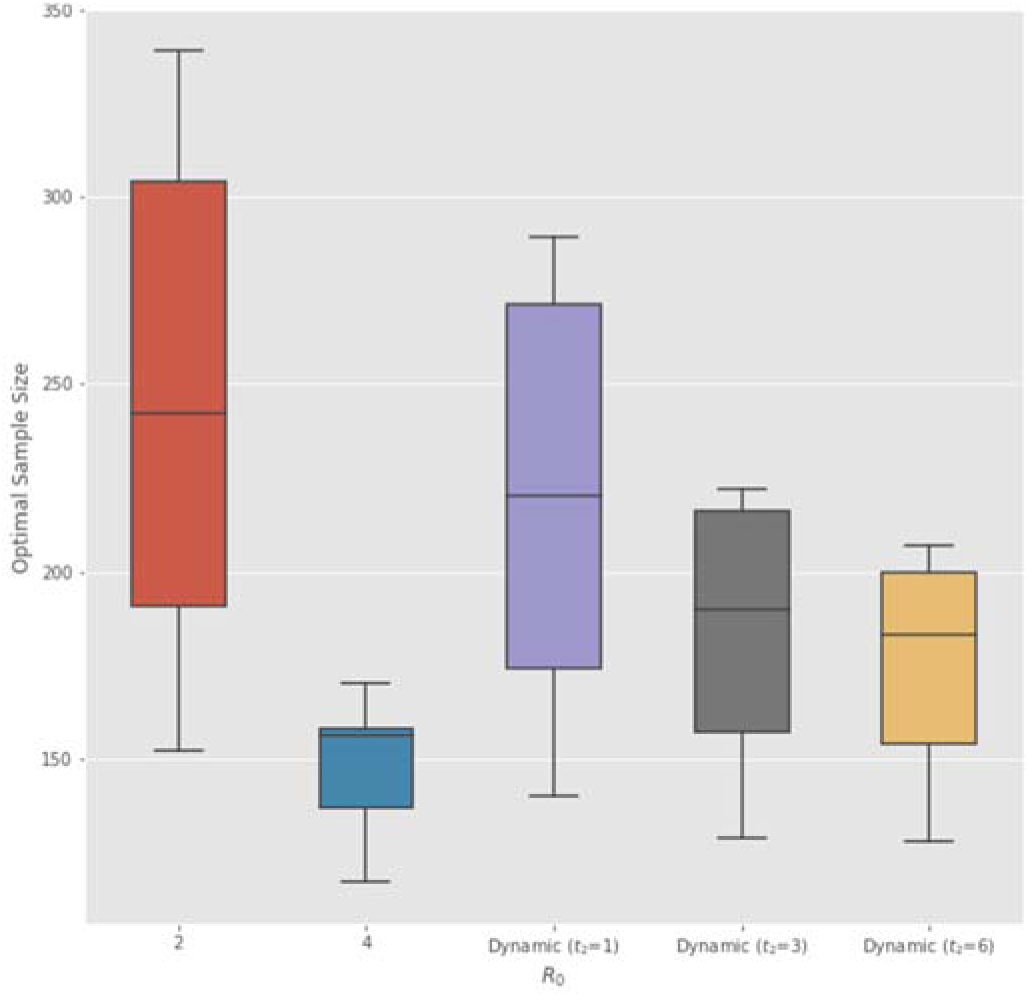
Sensitivity of optimal sample size (in each arm of non-adaptive Bayesian RCT) to the signal to noise ratio (SNR) ρ of treatment effect (Chaudhuri and Lo, 2018). From the upper to lower end of each box plot, the five-parameter summary corresponds to [50%, 75%, 100%, 125%, 150%] of the baseline value ρ = 0. 25 used in our analysis (Table 1).

**Figure 6.**
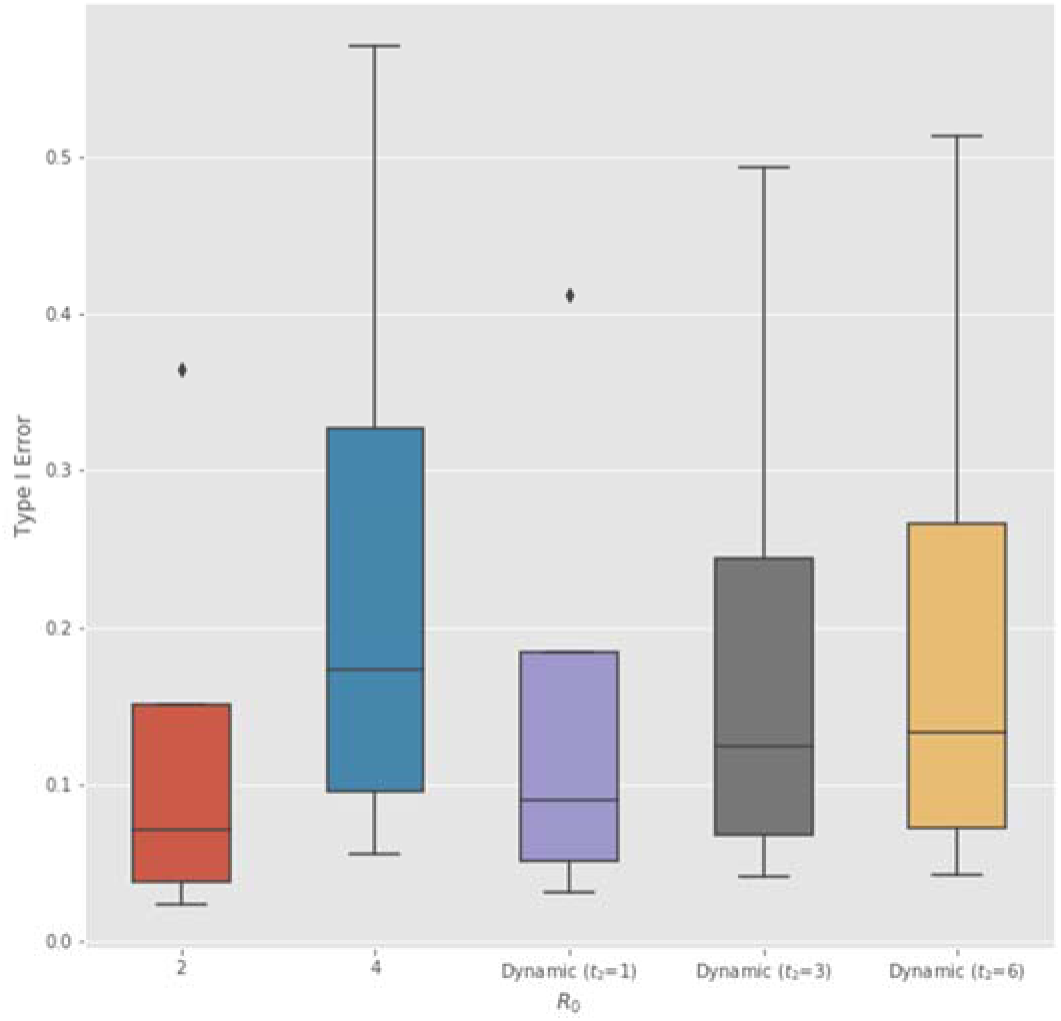
Sensitivity of optimal Type I error rate to the signal to noise ratio (SNR) ρ of treatment effect (Chaudhuri and Lo, 2018). From the upper to lower end of each box plot, the five-parameter summary corresponds to [50%, 75%, 100%, 125%, 150%] of the baseline value ρ = 0. 25 used in our analysis (Table 1).

**Figure 7.**
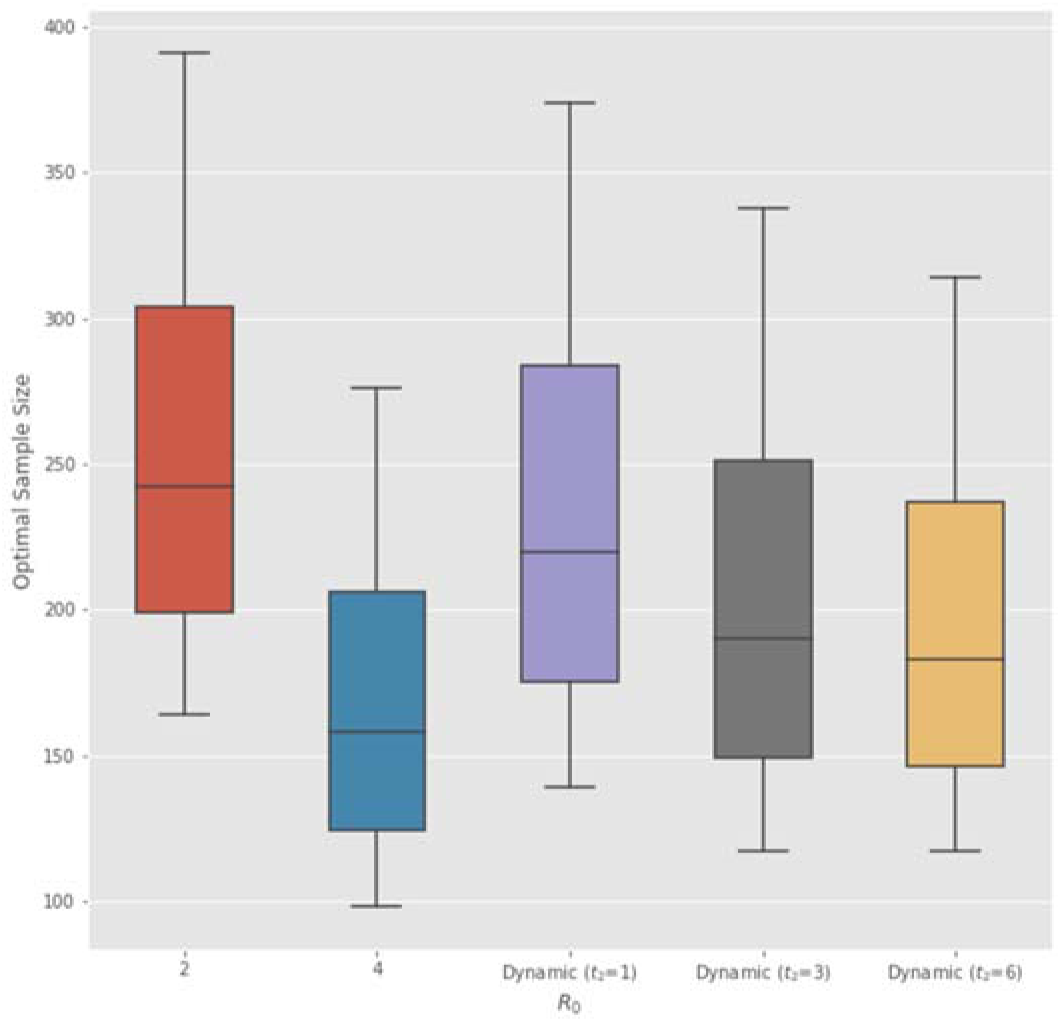
Sensitivity of optimal sample size (in each arm of non-adaptive Bayesian RCT) to the prior probability of having an ineffective treatment *p*_0_. From the lower to upper end of each box plot, the five-parameter summary corresponds to [70%, 85%, 100%, 115%, 125%] of the baseline value *p*_0_ = 77% used in our analysis (Table 1).

**Figure 8.**
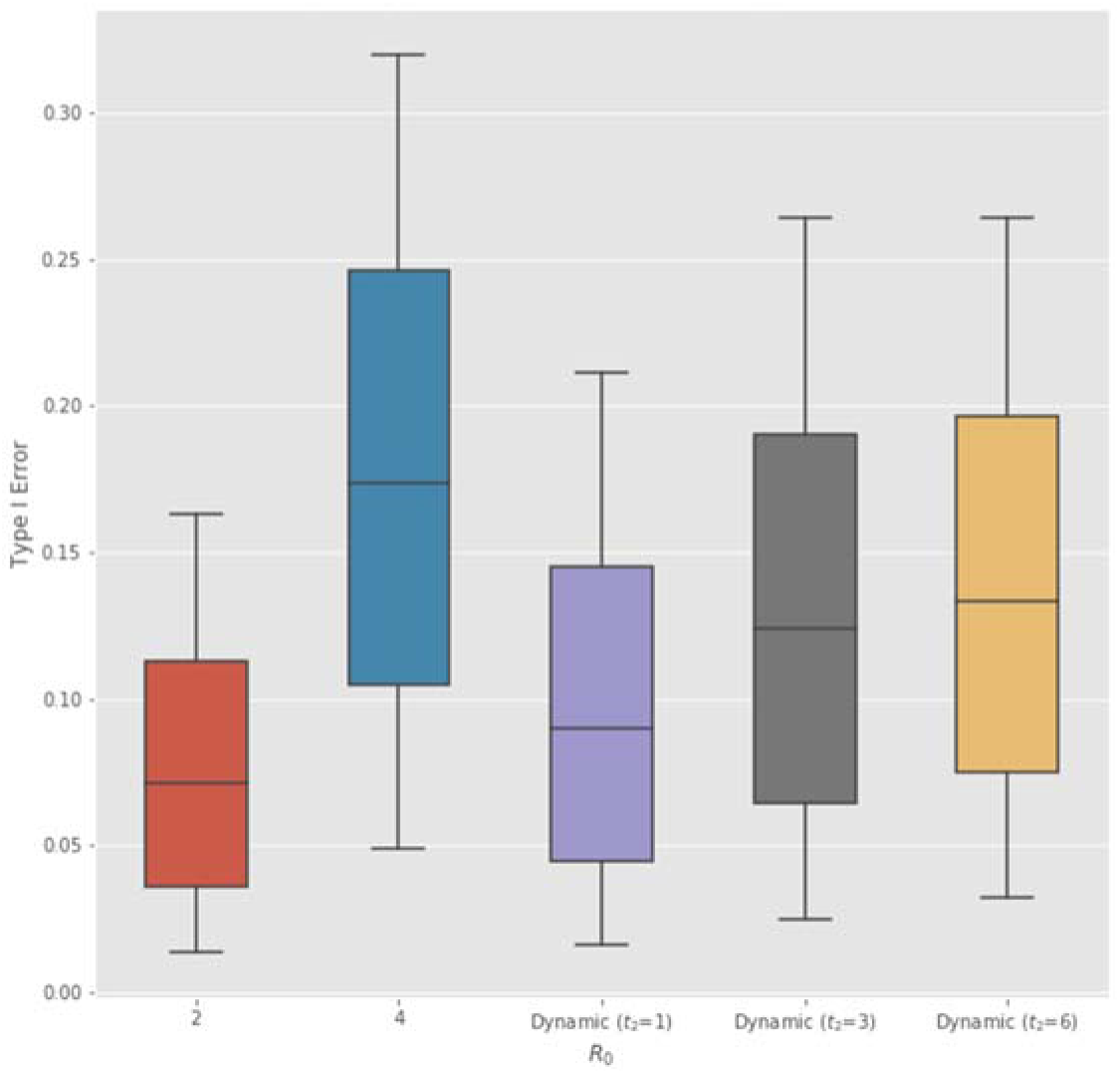
Sensitivity of optimal Type I error rate to the prior probability of having an ineffective treatment *p*_0_. From the upper to lower end of each box plot, the five-parameter summary corresponds to [70%, 85%, 100%, 115%, 125%] of the baseline value *p*_0_ = 77% used in our analysis (Table 1).

**Figure 9.**
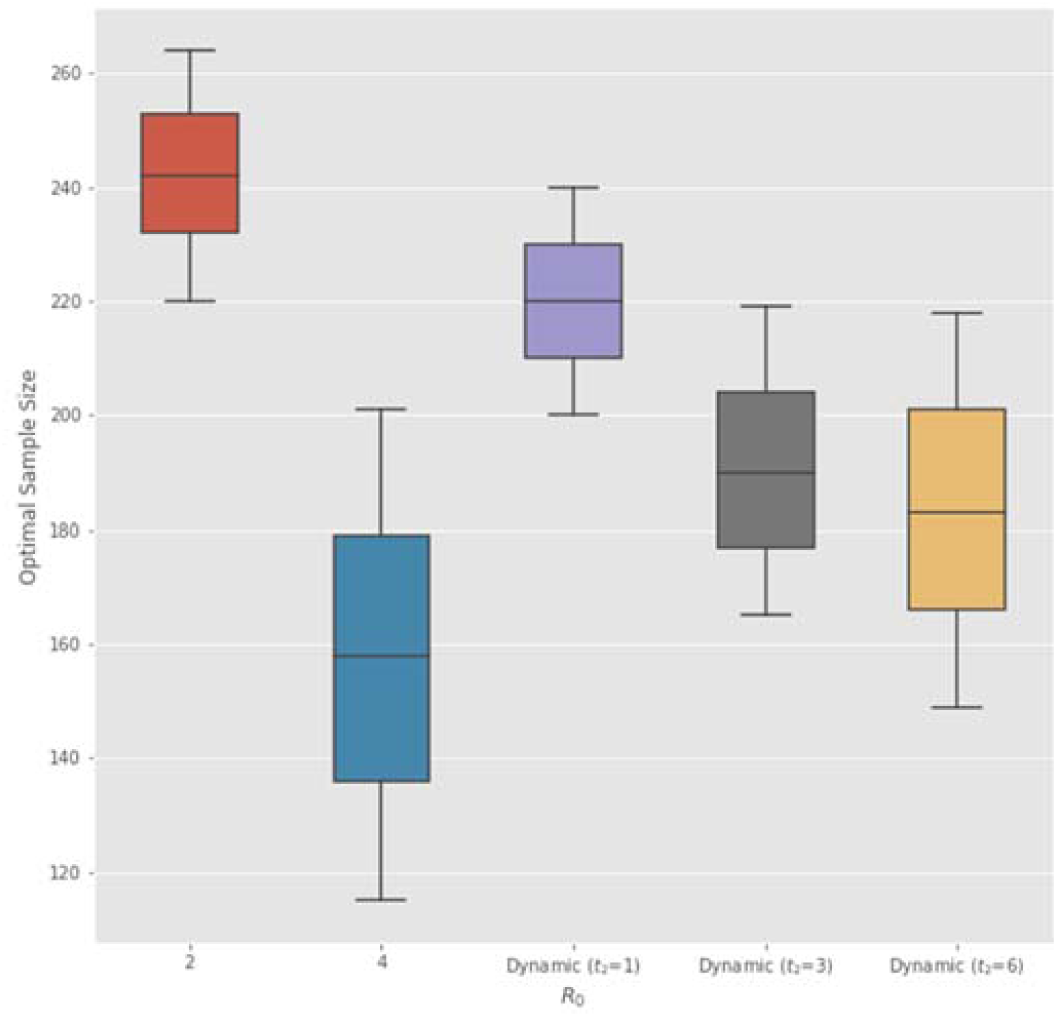
Sensitivity of optimal sample size (in each arm of non-adaptive Bayesian RCT) to the time to assess the efficacy of the non-vaccine anti-infective therapeutics Δ*t*. From the upper to lower end of each box plot, the five-parameter summary corresponds to [0%, 50%, 100%, 150%, 200%] of the baseline value Δ*t* = 1 week used in our analysis (Table 1).

**Figure 10.**
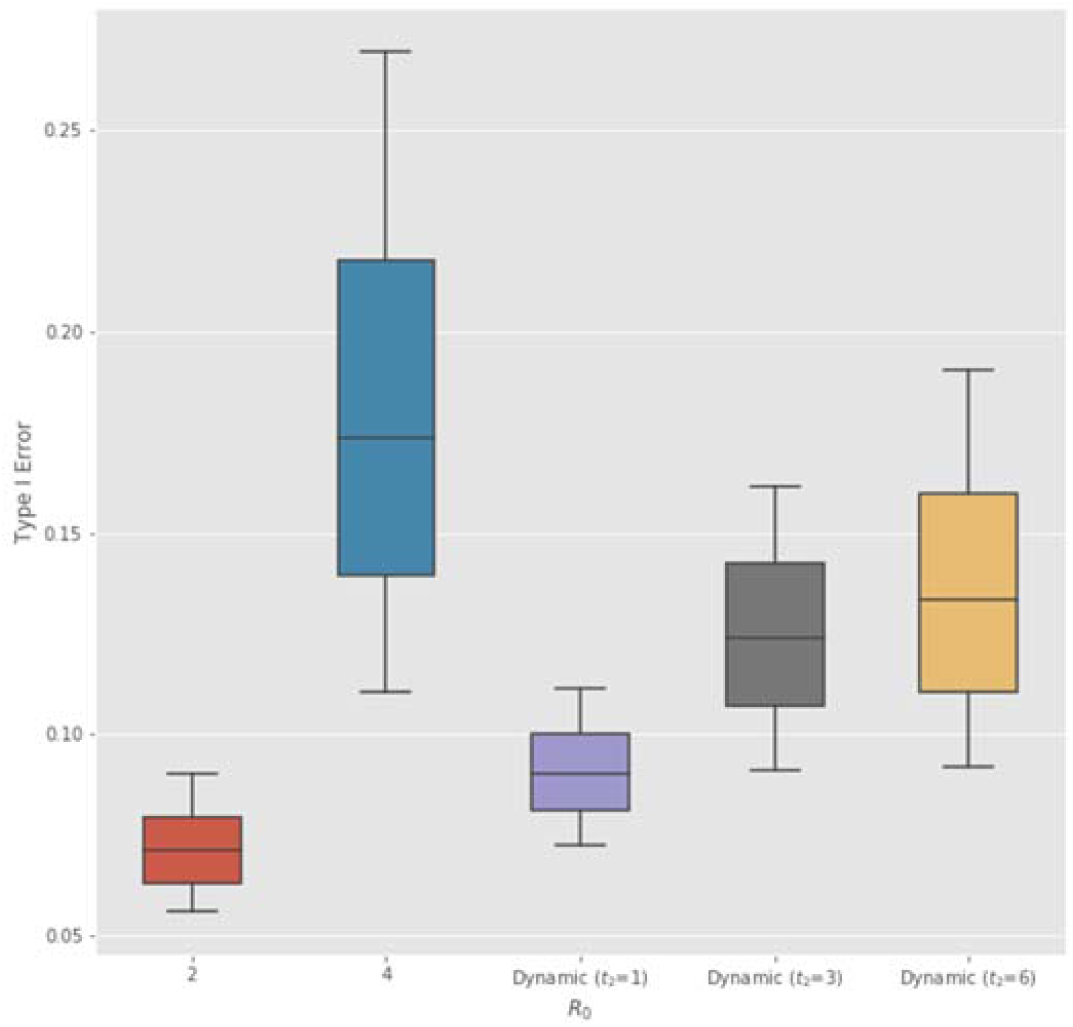
Sensitivity of optimal Type I error rate to the time to assess the efficacy of the anti-infective therapeutic Δ*t*. From the lower to upper end of each box plot, the five-parameter summary corresponds to [0%, 50%, 100%, 150%, 200%] of the baseline value Δ*t* = 1 week used in our analysis (Table 1).

**Figure 11.**
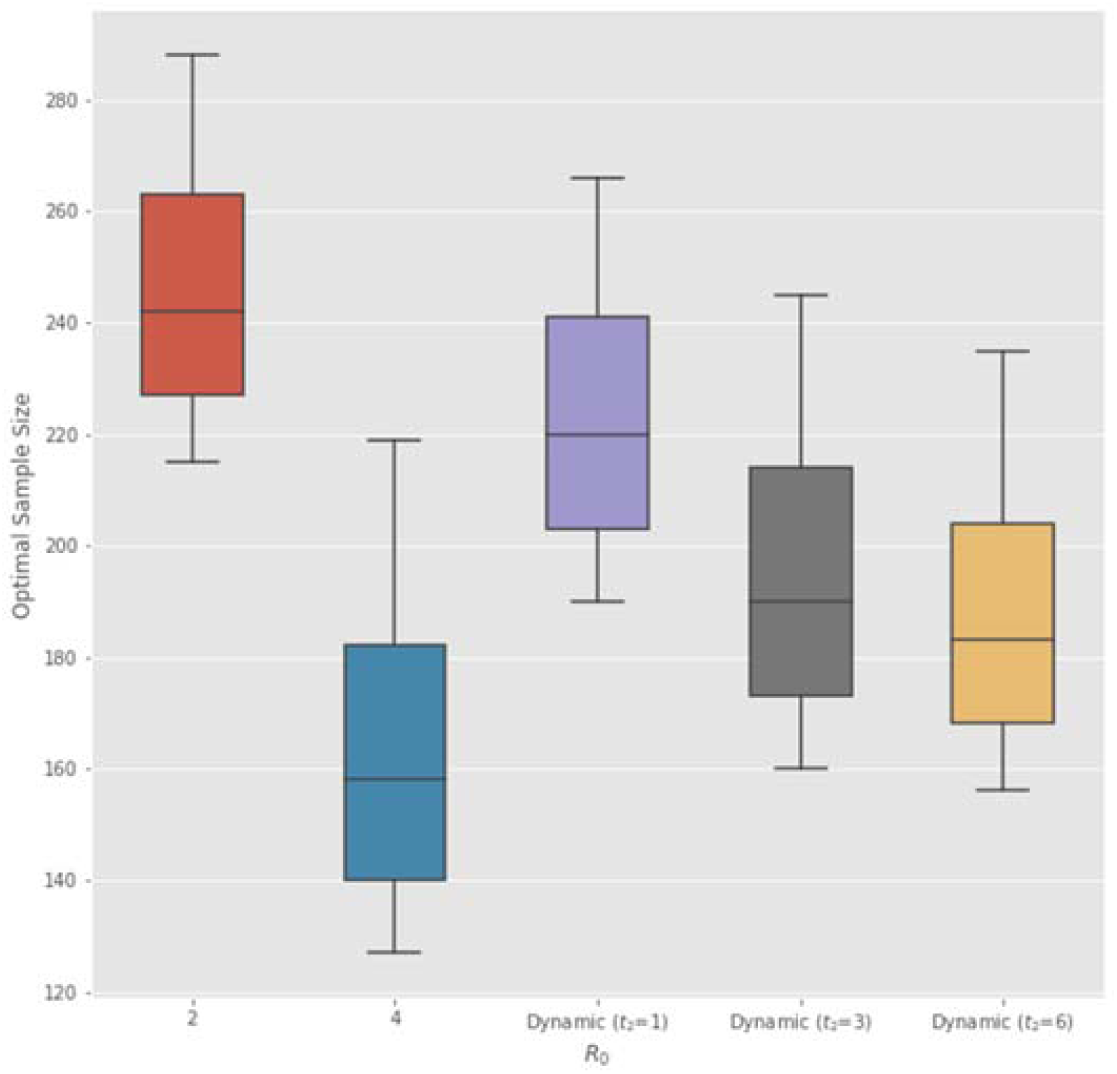
Sensitivity of optimal sample size (in each arm of non-adaptive Bayesian RCT) to the incubation rate *a* of the infectious disease. From the upper to lower end of each box plot, the five-parameter summary corresponds to [50%, 75%, 100%, 125%, 150%] of the baseline value *a* = 1 week used in our analysis (Table 1).

**Figure 12.**
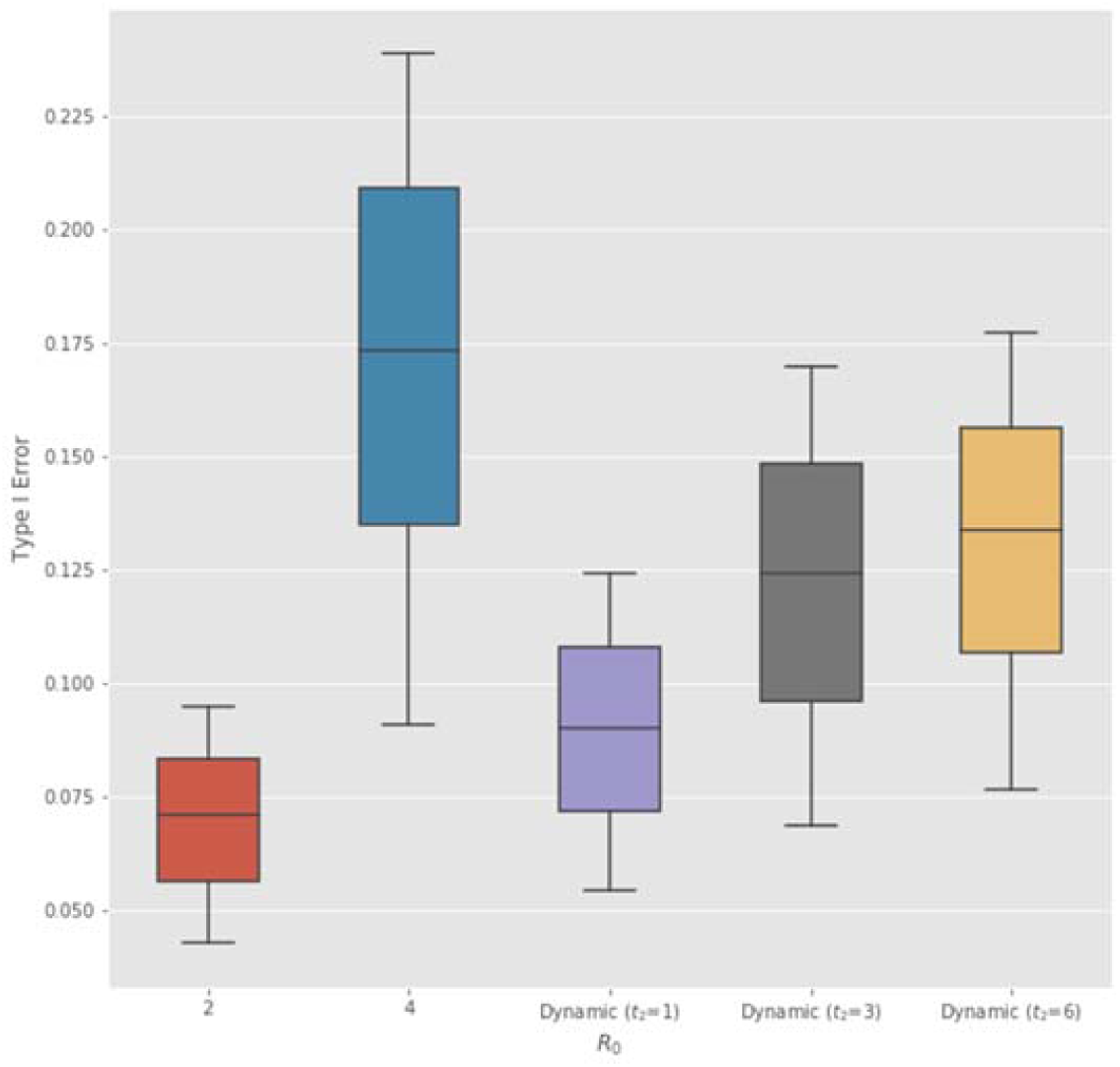
Sensitivity of optimal Type I error rate to the incubation rate *a* of the infectious disease. From the lower to upper end of each box plot, the five-parameter summary corresponds to [50%, 75%, 100%, 125%, 150%] of the baseline value *a* = 1 week used in our analysis (Table 1).

**Table 6.** Optimal sample size and Type I error rate *α* for Bayesian non-adaptive RCT on anti-infective therapeutics with *R*_0_ (basic reproduction number) close to 1. µ denotes the disease morality, and *I*_0_ the proportion of initial infected subjects. Sample size denotes the number of subjects enrolled in each arm of the RCT.

| Disease | $R_0$ | $I_0$ | Sample Size | $\alpha$ % | Power % |
| --- | --- | --- | --- | --- | --- |
| COVID-19 | 1.25 | 0.1% | 185 | 13.1 | 90 |
|  | 1.5 | 0.1% | 239 | 7.3 | 90 |
|  | 1.75 | 0.1% | 250 | 6.5 | 90 |
|  | 2 | 0.1% | 242 | 7.1 | 90 |
|  | 4 | 0.1% | 158 | 17.3 | 90 |
|  | 1.25 | 0.01% | 233 | 7.8 | 90 |
|  | 1.5 | 0.01% | 340 | 2.4 | 90 |
|  | 1.75 | 0.01% | 395 | 1.3 | 90 |
|  | 2 | 0.01% | 399 | 1.2 | 90 |
|  | 4 | 0.01% | 274 | 5.0 | 90 |
| SARS | 1.25 | 0.1% | 69 | 42.6 | 90 |
|  | 1.5 | 0.1% | 140 | 20.9 | 90 |
|  | 1.75 | 0.1% | 162 | 16.6 | 90 |
|  | 2 | 0.1% | 164 | 16.3 | 90 |
|  | 4 | 0.1% | 112 | 27.8 | 90 |
| MERS | 1.25 | 0.1% | 6 | 80.2 | 90 |
|  | 1.5 | 0.1% | 51 | 50.8 | 90 |
|  | 1.75 | 0.1% | 80 | 38.2 | 90 |
|  | 2 | 0.1% | 63 | 45.2 | 90 |
|  | 4 | 0.1% | 60 | 46.5 | 90 |

**Table 7.** Simulation results of Bayesian adaptive RCT on non-vaccine anti-infective therapeutics obtained from 10,000 Monte Carlo runs and assuming *L*_*D*_ = 10. *R*_0_ denotes the basic reproduction number, µ the disease morality, and *I*_0_ the proportion of initial infected subjects. Sample size refers to the number of subjects enrolled in each arm of the RCT. SD denotes standard deviation, and IQR the interquartile range about the median. *R*_0_(t) denotes dynamic transmission model with half-life *t*_2_ = 3 weeks and window length τ = 1 week.

| Epidemic Parameters |  |  | Non-Adaptive |  | Power % | Adaptive (100,000 Monte Carlo Runs) |  |  |  |  |  |
| --- | --- | --- | --- | --- | --- | --- | --- | --- | --- | --- | --- |
| | | | | | | Sample Size (H=0) | | Sample Size (H=1) | | $\alpha$ % | Power % |
| $R_0$ | $\mu$ | $I_0$ | Sample Size | $\alpha$ % | | Mean (SD) | Median (IQR) | Mean (SD) | Median (IQR) | | |
| 2 | COVID-19 | 0.1% | 281 | 4.7 | 90 | 141 (113) | 107 (63, 182) | 175 (121) | 142 (90, 226) | 3.7 | 91.3 |
| 4 | COVID-19 | 0.1% | 176 | 14.4 | 90 | 119 (86) | 96 (59, 153) | 108 (85) | 83 (48, 139) | 11.9 | 92.3 |
| $R_0(t)$ | COVID-19 | 0.1% | 213 | 9.7 | 90 | 128 (95) | 101 (61, 165) | 130 (97) | 53 (29, 98) | 7.7 | 91.9 |
| 2 | COVID-19 | 0.01% | 433 | 0.8 | 90 | 150 (128) | 109 (64, 191) | 272 (166) | 223 (156, 348) | 0.6 | 91.1 |
| 4 | COVID-19 | 0.01% | 290 | 4.2 | 90 | 143 (113) | 108 (63, 185) | 177 (119) | 145 (92, 229) | 3.3 | 91.1 |
| $R_0(t)$ | COVID-19 | 0.01% | 345 | 2.3 | 90 | 146 (118) | 111 (65, 188) | 217 (119) | 181 (117, 282) | 1.9 | 91.0 |
| 2 | SARS | 0.1% | 262 | 5.7 | 90 | 138 (107) | 107 (63, 179) | 161 (115) | 130 (81, 207) | 4.6 | 91.4 |
| 4 | SARS | 0.1% | 167 | 15.8 | 90 | 118 (86) | 94 (57, 154) | 102 (82) | 77 (46, 133) | 13.6 | 92.3 |
| $R_0(t)$ | SARS | 0.1% | 194 | 11.9 | 90 | 126 (93) | 100 (60, 165) | 118 (90) | 92 (55, 154) | 9.7 | 92.5 |
| 2 | MERS | 0.1% | 227 | 8.4 | 90 | 130 (97) | 102 (62, 167) | 140 (101) | 112 (68, 181) | 7.4 | 91.6 |
| 4 | MERS | 0.1% | 149 | 19.0 | 90 | 110 (78) | 89 (55, 142) | 93 (78) | 69 (39, 122) | 16.1 | 92.7 |
| $R_0(t)$ | MERS | 0.1% | 163 | 16.5 | 90 | 116 (84) | 93 (56, 151) | 101 (82) | 78 (45, 121) | 13.7 | 92.2 |

**Table 8.** Simulation results of Bayesian adaptive RCT on vaccines obtained from 10,000 Monte Carlo runs and assuming *L*_*D*_ = 10. *R*_0_ denotes the basic reproduction number, µ the disease morality, and *I*_0_ the proportion of initial infected subjects. Sample size refers to the number of subjects enrolled in each arm of the RCT. SD denotes standard deviation, and IQR the interquartile range about the median. *R*_0_ (t) denotes dynamic transmission model with half-life *t*_2_ = 3 weeks and window length τ = 1 week.

| Epidemic Parameters |  |  | Non-Adaptive |  |  | Adaptive (100,000 Monte Carlo Runs) |  |  |  |  |  |
| --- | --- | --- | --- | --- | --- | --- | --- | --- | --- | --- | --- |
| | | | | | | Sample Size (H=0) | | Sample Size (H=1) | | $\alpha$ % | Power % |
| $R_0$ | $\mu$ | $I_0$ | Sample Size | $\alpha$ % | Power % | Mean (SD) | Median (IQR) | Mean (SD) | Median (IQR) | | |
| 2 | COVID-19 | 0.1% | 221 | 8.9 | 90 | 130 (98) | 103 (62, 167) | 136 (100) | 109 (66, 176) | 7.5 | 91.9 |
| 4 | COVID-19 | 0.1% | 129 | 23.4 | 90 | 105 (78) | 84 (51, 135) | 80 (70) | 58 (33, 105) | 19.2 | 92.2 |
| $R_0(t)$ | COVID-19 | 0.1% | 151 | 18.7 | 90 | 114 (83) | 92 (56, 149) | 94 (78) | 69 (40, 123) | 15.4 | 92.4 |
| 2 | COVID-19 | 0.01% | 377 | 1.6 | 90 | 148 (126) | 110 (64, 187) | 236 (150) | 200 (130, 300) | 1.2 | 90.8 |
| 4 | COVID-19 | 0.01% | 247 | 6.7 | 90 | 135 (105) | 103 (63, 175) | 150 (106) | 121 (75, 196) | 5.4 | 91.3 |
| $R_0(t)$ | COVID-19 | 0.01% | 281 | 4.6 | 90 | 139 (109) | 107 (62, 181) | 170 (116) | 139 (88, 220) | 3.7 | 91.4 |
| 2 | SARS | 0.1% | 201 | 11.0 | 90 | 127 (94) | 100 (61, 165) | 122 (91) | 96 (58, 159) | 9.9 | 91.9 |
| 4 | SARS | 0.1% | 120 | 25.6 | 90 | 101 (74) | 81 (49, 131) | 76 (68) | 55 (30, 100) | 20.9 | 93.4 |
| $R_0(t)$ | SARS | 0.1% | 134 | 22.2 | 90 | 106 (76) | 86 (52, 139) | 83 (72) | 60 (33, 108) | 18.3 | 92.5 |
| 2 | MERS | 0.1% | 166 | 16.0 | 90 | 116 (84) | 92 (56, 152) | 102 (84) | 77 (45, 132) | 13.5 | 92.2 |
| 4 | MERS | 0.1% | 103 | 30.4 | 90 | 93 (70) | 75 (44, 122) | 67 (63) | 46 (24, 88) | 25.7 | 93.5 |
| $R_0(t)$ | MERS | 0.1% | 105 | 29.8 | 90 | 96 (71) | 77 (45, 125) | 68 (62) | 48 (25, 91) | 25.4 | 93.3 |

**Figure 13.**
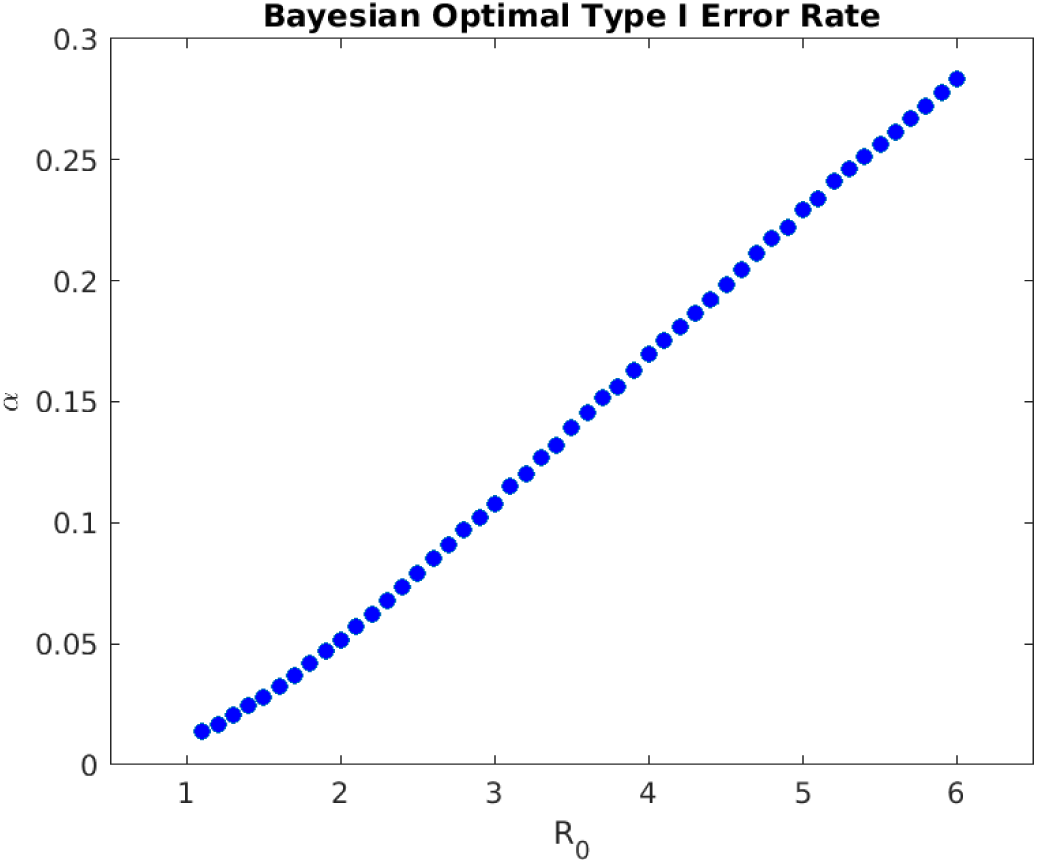
Optimal Type I error rate *α* of non-adaptive Bayesian RCT monotonically increases with the basic reproduction number *R*_0_ (assuming *I*_0_ = 0. 1%, *L*_*D*_ = 100 and disease mortality of COVID-19) if we define the loss of making a Type I error as the absolute risk of being susceptible *S*(*t*)*NL*_*S*_. This alternative definition is not very realistic. For an epidemic with *R*_0_ < 2, the loss of Type I error converges to a large positive value *S*(*T*)*NL*_*S*_ as time approaches the end of the epidemic outbreak. However, at the end of the outbreak, there are no more infected patients and thus no susceptible subjects. Therefore, the loss of Type I error should approach zero as *t* → *T*. This is the case for the excess risk of susceptibility (*S*(*t*) - *S*(*T*))*NL*_*S*_ but not for the absolute risk of susceptibility *S*(*t*)*NL*_*S*_.

